# Knowledge Graph-Guided Identification of Multiple Sclerosis and Therapeutic Trend Analysis: Real-World Evidence from Two Large Healthcare Systems

**DOI:** 10.1101/2025.11.12.25340116

**Authors:** Ziming Gan, Wen Zhu, Weijing Tang, Sara Morini Sweet, Michele Morris, Yunqing Han, Chenyi Chen, Junwei Lu, Emily Song, Mohammed Moro, Shyam Visweswaran, Tianrun Cai, Tanuja Chitnis, Tianxi Cai, Zongqi Xia

## Abstract

**Background:** The multiple sclerosis (MS) therapeutic landscape has evolved over time.

**Objective:** We conducted a knowledge graph-guided analysis of MS-specific disease-modifying therapy (DMT) prescription trends using longitudinal real-world clinical data.

**Methods:** We utilized registry-linked electronic health records (EHR) data from two large independent healthcare systems encompassing both academic and community practices (2004-2022). We applied a novel and efficient Knowledge-driven Online Multimodal Automated Phenotyping (KOMAP) algorithm to identify patients diagnosed with MS and evaluated algorithm performance against chart-reviewed and registry-recorded diagnosis labels. To assess temporal trends in DMT prescriptions, we combined the two cohorts and constructed time-varying temporal knowledge graphs using the patient-level EHR data segmented by calendar year. For each year, we analyzed co-occurrence patterns between DMTs and MS diagnosis by using Shifted Positive Pointwise Mutual Information transformation and singular value decomposition to generate embeddings. We computed patient-wise cosine similarities and confidence intervals.

**Results:** The phenotyping algorithm achieved robust performance in predicting MS diagnosis (AUROC: MGB=0.994, UPMC=0.922), identifying 29,169 MS patients in the combined dataset. Among commonly used standard-effectiveness DMTs, prescriptions for interferon-beta (slope=-0.018±0.011, p<0.001) and glatiramer acetate (slope=-0.013±0.012, p=0.026) and fumarates (slope=-0.031±0.010, p<0.001) declined after 2013. Use of S1P receptor modulators (slope=-0.026±0.016, p=0.005) declined after 2015. Among commonly used higher-effectiveness DMTs, B-cell depletion therapies (slope=0.051±0.027, p<0.001), particularly ocrelizumab (slope=0.020±0.016, p<0.001), showed a marked increase since 2017. Natalizumab usage peaked in 2012 (slope_pre-2012_=0.063±0.012, p_pre-2012_<0.001; slope_post-2012_=-0.027±0.008, p_post-2012_<0.001). Other DMT classes such as cell proliferation inhibitors, chemotherapy agents, and purine blockers, showed low usage during follow-up.

**Conclusion:** Real-world evidence from two large EHR-based MS cohorts highlights distinct temporal shifts in the MS therapeutic landscape toward higher-effectiveness DMTs, particularly B-cell depletion therapy.

**Key Messages:**

1. Accurate identification of patients diagnosed with multiple sclerosis (MS) from real-world clinical data is essential for tracking longitudinal prescription patterns at scale and understanding the evolution of the MS therapeutic landscape.
2. Leveraging electronic health records (EHR) data, our knowledge graph-guided unsupervised algorithm accurately and efficiently identified MS patients from two large, independent healthcare systems.
3. Temporal analysis using knowledge graph-guided methods revealed major shifts in MS-related disease-modifying therapy (DMT) prescriptions, including a decline in early injectable use and increased adoption of B-cell depletion therapies.
4. These findings confirm a growing preference for higher-effectiveness DMTs in MS and provide a scalable framework for evaluating long-term treatment patterns across healthcare systems.

## Introduction

Multiple Sclerosis (MS) is a chronic autoimmune disease characterized by inflammatory demyelination and progressive neurodegeneration in the central nervous system, leading to neurological impairments.^1^ Affecting millions of people worldwide, MS diminishes individual quality of life and creates disproportionally high societal healthcare burdens,^2^ partly driven by the cost of disease-modifying therapies (DMTs). Given the wide range of DMT options and variability in clinical practices, robust methods for examining long-term DMT prescription trends are essential. Such approaches can evaluate real-world adoption of clinical guidelines, identify gaps in patient access to DMTs, inform clinicians of emerging therapeutic patterns across demographic or clinical subgroups, guide healthcare systems in resource planning (*e.g.,* building infusion centers), assist payors in coverage decisions for expanding DMT options, and ultimately contribute to the long-term real-world evidence of DMT effectiveness, safety, and adherence.

Accurate classification of MS cases through robust phenotyping using *routine* clinical data from electronic health records (EHR) is essential for generating large-scale real-world evidence, given the spectrum of MS presentation and variability in clinical documentation. Traditional rule-based algorithms often rely on limited structured data, such as diagnostic codes or medication history, which can result in misclassification and reduced generalizability across healthcare systems.^3–6^ KOMAP (Knowledge driven Online Multimodal Automated Phenotyping) addresses these limitations by integrating codified data (*e.g.*, ICD codes, medication records) with unstructured data (*e.g.*, clinical notes) through ontological mapping (to common features) and machine learning.^7^ By leveraging multimodal features and preserving clinical context, KOMAP enables robust and scalable MS phenotyping across diverse EHR systems, facilitating accurate identification of patient cohorts for observational research and precision medicine efforts.

Over the years, the landscape of MS treatment has evolved significantly, with new drugs being introduced and their effectiveness re-evaluated as more clinical data become available.^8–11^ The relationships between drugs and MS are dynamic, shaped by factors such as drug development, regulatory approvals, clinical guidelines, and emerging evidence from clinical trials and observational studies. This temporal variability highlights the importance of examining how these relationships change over time. A comprehensive, time-varying knowledge graph (KG) can support the systematic analysis of these evolving connections, offering a powerful tool to map the MS profiles alongside the corresponding therapeutic landscape.^12,13^ Such a KG can not only reveal historical trends but also support predictive modeling of future drug-disease interactions, ultimately contributing to a more informed and adaptive approach to MS management. While several studies have focused on constructing temporal KGs for EHR data; however, these works primarily emphasize tracking the dynamic progression of individual patients rather than capturing broader population-level developments.^14^

In this study, we aim to construct a temporal KG of DMT usage among KOMAP-identified MS patients from two independent cohorts, capturing shifts in drug-disease relationships over the years. By analyzing these temporal patterns, we seek to uncover the underlying trends, thereby informing future investigations into the factors driving the evolving treatment paradigms for MS and supporting the development of targeted therapeutic interventions to advance precision medicine.

## Methods

### Ethics Approval

The institutional review boards of the University of Pittsburgh (STUDY20070274 and STUDY21030127) approved the study protocols. The use of de-identified clinical data was deemed exempt.

### Study Design and Datasets

Figure 1 illustrates the overall study design. We obtained EHR data from two large independent healthcare systems, UPMC (Pittsburgh, PA) and Mass General Brigham (MGB: Boston, MA), each comprising both academic and community practices as well as inpatient and outpatient records from multiple EHR platforms (*e.g.*, Epic, Cerner). Both healthcare systems provided *codified data* from 2004-2022, encompassing demographics (*e.g.*, age, sex, race, ethnicity), diagnosis and procedure codes (*e.g.*, International Classification of Disease [ICD] code, Current Procedural Terminology [CPT] code), healthcare utilization metrics (*e.g.*, total number of ICD codes and encounter notes), medication history (e.g., RxNorm electronic prescriptions), and laboratory results (*e.g.*, Logical Observation Identifiers Names and Codes [LOINC]). Further, *narrative text data* from 2011-2022 were available, comprising free-text documentation from clinical encounters (*e.g.*, physician office visit notes, discharge summaries), which were transformed into structured data using natural language processing (NLP). We constructed an MS data mart within each healthcare system consisting of patients with at least one ICD diagnosis code for MS as an *initial* step to enhance sensitivity.

**Figure 1.**
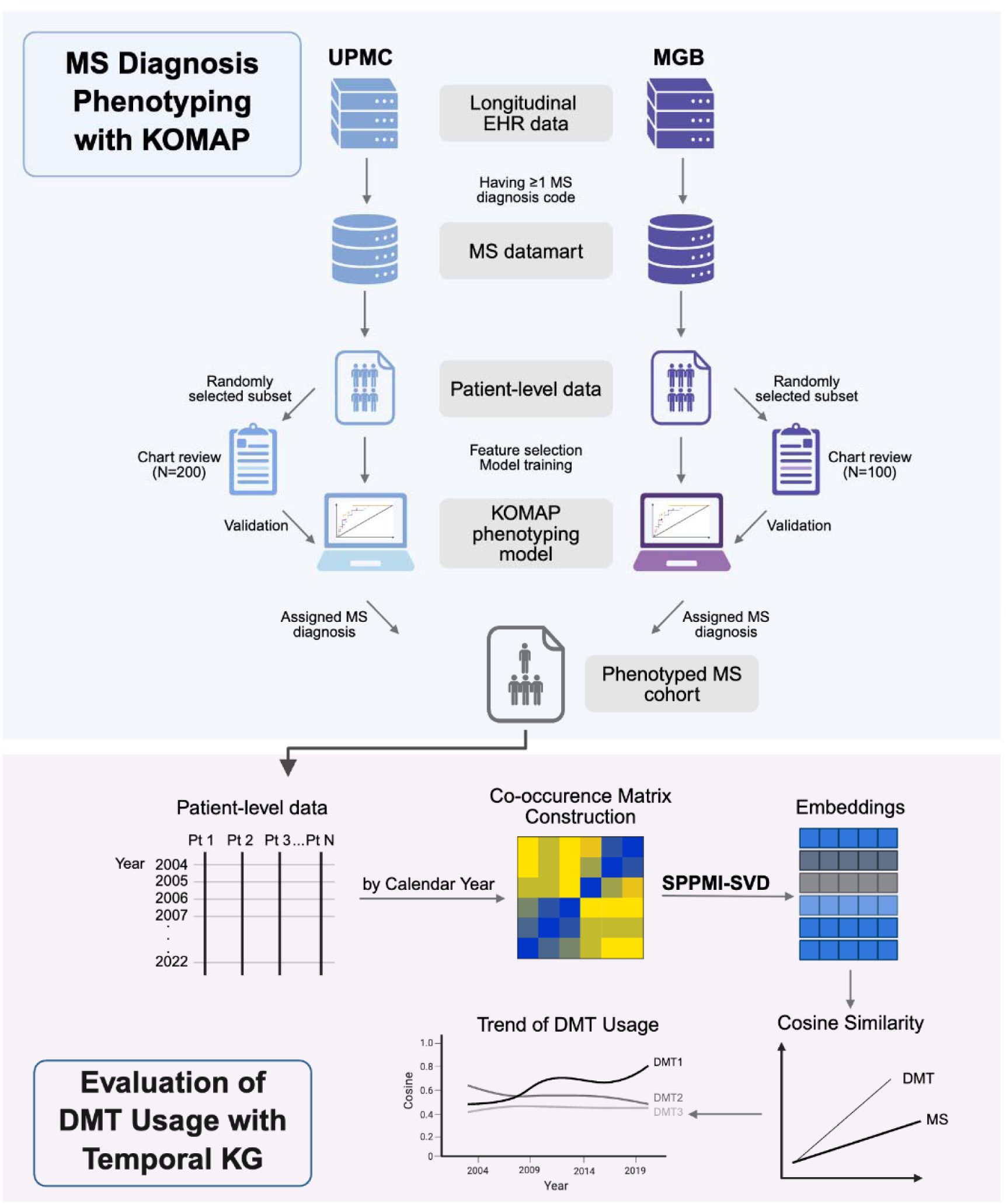
Overall study design. Top panel: MS phenotyping with KOMAP to identify MS cases. Bottom panel: construction of temporal knowledge graphs from EHR data. Patient records were filtered and partitioned by calendar year. For each year, a co-occurrence matrix of EHR codes was computed, followed by Shifted Positive Pointwise Mutual Information (SPPMI) transformation and singular value decomposition (SVD) to generate embeddings. Cosine similarity matrices derived from these embeddings represented relationships among codes. Temporal trends were analyzed using smoothed similarity estimates, enabling year-over-year comparison and interpretation of evolving clinical patterns.

### Dimensionality Reduction of Codified and Narrative Features

We standardized the codified data to ensure consistent representation of clinical concepts. Specifically, we mapped all ICD-9 and ICD-10 diagnosis codes to PheCodes,^15^ laboratory tests to LOINC,^16^ and prescriptions to ingredient-level RxNorm codes.^17^ Procedure codes (CPT) were grouped using the Clinical Classifications Software for Services and Procedures. To extract standardized concepts from narrative clinical text, we applied a previously validated NLP pipeline, *i.e.,* Narrative Information Linear Extraction (NILE).^18^ This pipeline generates concept unique identifiers (CUIs) based on the Unified Medical Language System (UMLS).^19^

### Assigning and Validating MS Diagnosis

We applied a novel phenotyping algorithm, KOMAP, to determine MS diagnosis status (MS vs non-MS) for all patients in the initial MS data mart.^7^ KOMAP is an unsupervised, two-step algorithm designed to extract accurate diagnostic labels from EHR data. In the first step, KOMAP uses the Online Narrative and Codified feature Search engine (*i.e.,* ONCE), powered by multi-source KG representation learning, to identify informative codified and narrative features related to MS. In the second step, the algorithm is trained using the features selected by ONCE. These features include strong surrogate indicators of MS diagnosis (*e.g.*, the primary MS PheCode from structured data and the main MS CUI from narrative data) along with a measure of healthcare utilization defined by the total number of ICD codes and clinical encounters. We validated KOMAP-assigned MS diagnosis against chart-reviewed labels (annotated by trained domain experts) in a randomly selected subset of patients (n=200 in UPMC, n=100 in MGB).

We evaluated the performance of different feature sets (*i.e.,* main MS PheCode, main MS CUI, codified features alone [“codified”], and combined codified plus NLP-derived features [“both”]) across two healthcare systems, using models with main PheCode and main CUI as benchmark. Performance was assessed using area under the receiver operating characteristic curve (AUROC), area under the precision-recall curve (AUPRC), and threshold-based sensitivity, positive predictive value (PPV), and negative predictive value (NPV) at predefined specificity levels (0.97, 0.95, and 0.90).

### Temporal KG of MS-specific DMT Usage

To enhance the generalizability and statistical power of our findings, we combined codified EHR data from UPMC and MGB. Merging these cohorts enabled the inclusion of a broader and more diverse patient population across geographic regions, demographic groups, and clinical practice patterns. This integration minimized site-specific biases and strengthened the robustness of identified trends and associations. Further, leveraging data from two distinct healthcare systems allowed for cross-system validation, increasing confidence in the reproducibility and external validity of our results.

### Knowledge Graph Construction

To construct the temporal KGs, we began by dividing the codified EHR data of KOMAP-classified MS patients from UPMC and MGB according to the calendar year in which each record was documented. For each year, a KG is created using the EHR data recorded during that period. A widely used approach for constructing KGs involves analyzing the occurrence and co-occurrence patterns of EHR codes. To extract meaningful relationships from the co-occurrence matrix, we applied the Shifted Positive Pointwise Mutual Information transformation and singular value decomposition (SPPMI-SVD) algorithm.^20^

Specifically, for a given year *t*, we constructed the co-occurrence matrix of codes, denoted as 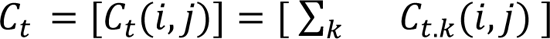, where *C_t,k_(i,j)* represented the total co-occurrence of the *i*th and *j*th codes within 30-day moving windows in the *k*th patient’s health record during calendar year *t*. The *(i,j)*th entry of the Shifted Positive Pointwise Mutual Information (SPPMI) matrix for calendar year *t* was then calculated as 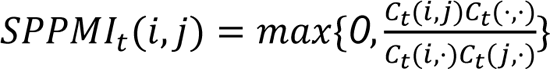, where *C_t_(i,.)* was the row sum of *C_t_* and *C_t_(.,.)* was the total sum of all entries in *C_t_*.

The SPPMI-SVD algorithm generated embeddings for the th calendar year by performing singular value decomposition (SVD) on the SPPMI matrix, represented as 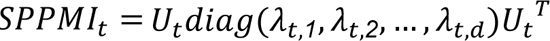. We then obtained the embedding for each node in year *t* as 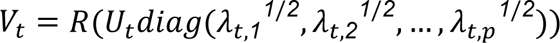, where *p* was the embedding dimension and *R* was the unit-length normalization operator that standardized each row to unit length. Using these embeddings, we computed the cosine similarity matrix for year *t* as *V_t_V_t_T*. To analyze temporal trends, we fit a Locally Estimated Scatterplot Smoothing (LOESS) function for each feature pair *(i,j)*, producing a smoothed estimator.

To construct confidence intervals for these estimators, we replicated the above method for each individual patient. Instead of using *C_t_(i,j)*, we used *C_t,k_(i,j)* to compute the patient-specific cosine similarity matrix as *V_kt_V_kt_T* for the th patient. Finally, for each calendar year, we aggregated the patient-wise cosine similarity values to construct confidence intervals based on the standard error across patients. This approach ensured robust and interpretable temporal KG representations.

### Change-point Detection

After computing the yearly cosine similarity for each feature pair *(i,j)*, we fit a piecewise (segmented) regression model to characterize the temporal trend in their relationship.^21^ Specifically, the model assumed that the cosine similarity over time could be described by a single linear regression model or two linear segments joined at unknown breakpoints. For a model with a single breakpoint at year τ, the cosine similarity was modeled as: *y_t_ = β_0_ + β_1_t + ∈_t_* when *t* ≤ τ, and *y_t_ = β_0_ + β_1_t + β_2_(t−τ) + ∈_t_* when *t* > τ. The breakpoint τ and parameters (β_0_,β_1_,β_2_) were estimated by minimizing the residual sum of squares. By applying this model to each feature pair, we captured both the overall direction of the trend and any structural shifts over time. Based on the fitted models, we categorized the feature pairs into four groups: (1) pairs with no significant trend changes, (2) pairs exhibiting a significantly increasing trend, (3) pairs exhibiting a significantly decreasing trend, and (4) pairs with a significant change point indicating a shift in the trend.

## Results

### Patient Characteristics

The study cohort included a total of 29,169 patients, comprising 10,301 from UPMC and 18,868 from MGB (**Table 1**). The overall median age at diagnosis was 45.9 years (IQR: 36.4–56.8), with a higher median age in the UPMC cohort (47.4 years) compared to MGB (44.8 years). The median disease duration was 12.4 years (IQR: 7.0–12.7), shorter in UPMC (11.8 years) and longer in MGB (12.8 years). The majority of the cohort were women (74.3%), with similar sex distributions across sites. Most patients identified as White (89.4%), followed by Black (6.5%) and Asian (1.2%), with racial composition relatively consistent across sites. Overall, 54.5% of patients had ever received a DMT, with a higher rate in UPMC (69.4%) than in MGB (47.3%).

**Table 1.** Cohort Characteristics of KOMAP-identified MS Patients.

|  | **All** | **UPMC** | **MGB** |
| --- | --- | --- | --- |
| **N** | 29169 | 10301 | 18868 |
| **Age at Diagnosis [Median (IQR)]** | 45.9<br>(36.4-56.8) | 47.4<br>(37.4-56.5) | 44.8<br>(34.7-54.9) |
| **Disease Duration [Median (IQR)]** | 12.4<br>(7.0-12.7) | 11.8<br>(8.2-12.5) | 12.8<br>(6.8-19.3) |
| **Gender [n (%)]** |  |  |  |
| **Women** | 21671 (74.3) | 7676 (74.5) | 13995 (74.2) |
| **Men** | 7497 (25.7) | 2625 (25.5) | 4872 (25.8) |
| **Unknown** | 1 (0.0) | 0 (0.0) | 1 (0.0) |
| **Race [n (%)]** |  |  |  |
| **American Indian or Alaska Native** | 66 (0.2) | 14 (0.1) | 52 (0.3) |
| **Asian** | 341 (1.2) | 42 (0.4) | 299 (1.6) |
| **Black** | 1893 (6.5) | 821 (8.0) | 1072 (5.7) |
| **Native Hawaiian or Other Pacific Islander** | 5 (0.0) | 2 (0.0) | 3 (0.0) |
| **White** | 26072 (89.4) | 9271 (90.0) | 16801 (89.1) |
| **Unknown** | 792 (2.7) | 151 (1.5) | 641 (3.4) |
| **Ethnicity [n (%)]** |  |  |  |
| **Hispanic or Latino** | 424 (1.5) | 55 (0.5) | 369 (2.0) |
| **Not Hispanic or Latino** | 28309 (97.1) | 9811 (95.2) | 18498 (98.0) |
| **Unknown** | 436 (1.4) | 435 (4.3) | 1 (0.0) |
| **Ever on DMT [n (%)]** |  |  |  |
| **Yes** | 15905 (54.5) | 7114 (69.4) | 8791 (47.3) |
| **No** | 13264 (45.5) | 3187 (30.6) | 10077 (52.7) |

### MS Diagnosis Phenotyping Algorithm Performance

At UPMC, the combined codified plus NLP-derived narrative feature set achieved the best performance with an AUROC of 0.922 and an AUPRC of 0.966. It outperformed both benchmark models (main MS PheCodes: AUROC 0.854, AUPRC 0.941; main MS CUI: AUROC 0.798, AUPRC 0.898) as well as codified-only feature set (AUROC 0.912, AUPRC 0.963). Similarly, at MGB, the combined feature set yielded the best performance, with an AUROC of 0.994 and AUPRC of 0.940 (Figure 2A, **Supplementary Table 2**). Across all specificity thresholds, the combined feature set consistently achieved the highest sensitivity and NPV, while PPV remained high across all models and healthcare systems, frequently exceeding 0.95 (**Supplementary Table 2**). Accordingly, we deployed the model using the combined feature set at a 0.900 specificity threshold to phenotype MS diagnosis and classify patients with MS in both the UPMC and MGB cohorts (Figure 2B and C).

**Figure 2.**
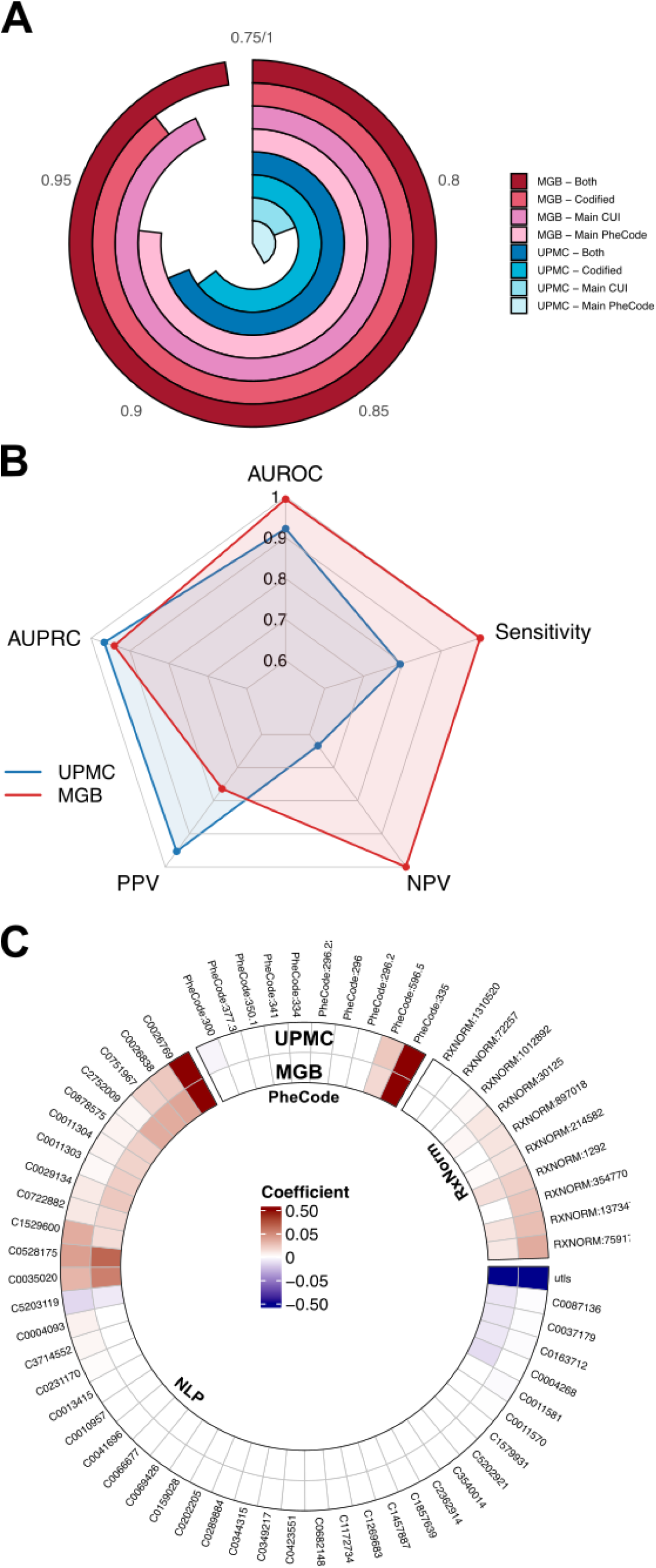
MS phenotyping model (KOMAP) performance and feature contributions. **A.** Circular plot showing the area under the receiver operating characteristic curve (AUROC) for KOMAP models trained with different feature sets (*i.e.,* main MS PheCode, main MS CUI, codified features alone [“codified”], and combined codified plus NLP-derived features [“both”]) across two independent healthcare systems (*i.e.,* UPMC and MGB). **B.** Radar plot comparing the KOMAP model performance metrics (*i.e.,* AUROC, area under the precision-recall curve [AUPRC], sensitivity, positive predictive value [PPV], and negative predictive value [NPV] between UPMC (blue) and MGB (red) at a pre-defined specificity threshold of 0.90, using the combined codified and narrative feature set for MS phenotyping. **C.** Circular bar plot displaying feature coefficients used in the KOMAP model at UPMC and MGB. Features are grouped by data type: PheCode (diagnoses), RxNorm (medications), and NLP-derived narrative concepts. Feature importance is color-coded by coefficient value (red = positive, blue = negative, white = neutral), with stronger contributions appearing as darker shades. Please refer to S-Table 1 for descriptions of the features.

### Time-varying Relationships between MS and DMTs

We next assessed temporal trends in MS therapies among KOMAP-classified MS patients using the temporal KG, highlighting dynamic shifts in the cosine similarity between MS and DMTs (*i.e.,* MS-specific therapies) as well as changes in DMT prescription frequency (Figure 3 and **Table 2**). It is important to note that prescription frequency does not necessarily indicate that the drug is primarily used to treat MS. Rather, cosine similarity between drug and MS provides a more direct measure of how specifically and frequently a drug is associated with MS treatment. This distinction underscores the value of cosine similarities as a focused metric for understanding the drug’s relationship to the disease.

**Figure 3.**
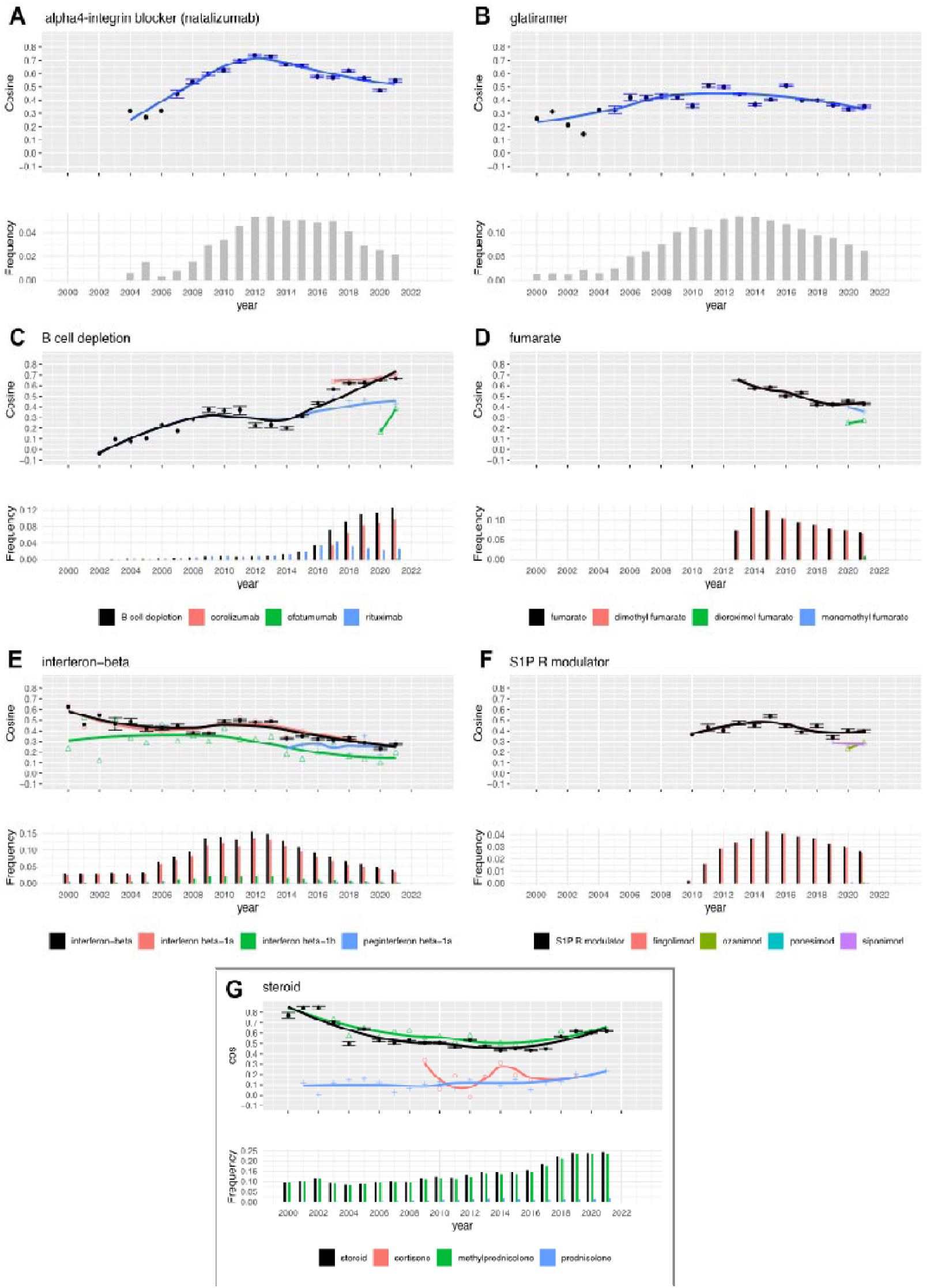
Trends of commonly prescribed MS-related DMT classes (2004-2022). **A.** α4-integrin blockers, **B.** B-cell depletion, **C**. fumarates, **D**. glatiramer acetate, **E.** interferon-beta, **F.** S1P receptor modulators, and **G**. steroids (included as a reference). The RxNorm code 214582 refers to the active ingredient glatiramer, including GA, Copaxone, and Glatopa. For each drug class, the top panel displays the dynamic cosine similarity with 95% confidence intervals. The bottom panel shows the DMT frequency, defined as the proportion of MS patients prescribed the target drug in each calendar year.

**Table 2.** Trends of common MS-related DMT classes between 2004 and 2022.

| DMT Class | Follow-up Period | Trend<sup>1</sup> | Change Point<sup>2</sup> | Before Change Point |  | After Change Point |  |
| --- | --- | --- | --- | --- | --- | --- | --- |
| | | | | $\beta$ (95% CI) | p | $\beta$ (95% CI) | p |
| Alpha4-integrin blocker | 2004-2022 | I-D | 2011 | 0.063<br>(0.050, 0.077) | <0.001 | -0.027<br>(-0.035, -0.019) | <0.001 |
| B cell depletion | 2002-2022 | I-I | 2014 | 0.022<br>(0.009, 0.035) | 0.002 | 0.051<br>(0.023, 0.078) | 0.001 |
| Fumarate | 2013-2022 | D-NA | 2020 | -0.028<br>(-0.042, -0.013) | 0.004 | -0.097<br>(-0.234, 0.041) | 0.135 |
| Glatiramer | 2000-2022 | I-D | 2011 | 0.022<br>(0.012, 0.032) | <0.001 | -0.013<br>(-0.025, -0.002) | 0.026 |
| Interferon-beta | 2000-2022 | NA-D | 2011 | -0.007<br>(-0.017, -0.002) | 0.124 | -0.019<br>(-0.030, -0.008) | 0.002 |
| S1P-R modulator | 2010-2022 | NA-D | 2014 | 0.022<br>(-0.011, 0.055) | 0.166 | -0.026<br>(-0.043, -0.010) | 0.005 |
| Steroid | 2000-2022 | D-I | 2013 | -0.027<br>(-0.037, -0.018) | <0.001 | 0.029<br>(0.011, 0.048) | 0.003 |

<sup>1</sup> **Trend:** the pattern of MS-related DMT prescription over time assessed using cosine similarity.
I-D, increasing trend before the change point, followed by a decreasing trend;
I-I, increasing throughout, at different rate before and after the change point;
D-I, decreasing trend, followed by an increasing trend;
D-NA, decreasing trend with no significant trend after the change point;
NA-D, no significant trend before the change point, followed by a decreasing trend.
<sup>2</sup> **Change point:** the year during which a statistically significant shift in the MS-related DMT prescription trend occurred.

We examined the major DMT mechanistic classes. For the alpha-4-integrin blocker (*i.e.,* natalizumab), cosine similarity increased steadily from 2004 [β (95% CI) = 0.063 (0.050, 0.077), *p*<.001], peaked around 2011, and then gradually declined [β (95% CI) = –0.027 (–0.035, – 0.019), *p*<.001], mirroring its prescription frequency trend, which also peaked around 2011–2012 (Figure 3A). Glatiramer acetate initially showed a moderate, stable increase in cosine similarity [β (95% CI) = 0.022 (0.012, 0.032), *p*<.001] until 2011, followed by a decline [β (95% CI) = –0.013 (–0.025, –0.002), *p*<.001], with frequency peaking in 2015 before decreasing (Figure 3B). B-cell depletion therapies (*i.e.,* ocrelizumab, ofatumumab, and rituximab) exhibited a sharp increase in cosine similarity beginning around 2014 [β (95% CI) = 0.051 (0.023, 0.078), *p*=.001], which accelerated following the introduction of ocrelizumab post-2017, paralleled by a continuous rise in prescription frequency (Figure 3C). In contrast, fumarates (*i.e.,* dimethyl fumarate, diroximel fumarate, and monomethyl fumarate) showed a modest downward trend in cosine similarity following the introduction of dimethyl fumarate in 2013 [β (95% CI) = –0.028 (–0.042, –0.013), *p*=.001], but exhibited a relatively flat trend after the introduction of diroximel fumarate and monomethyl fumarate around 2020, which matched the prescription frequency pattern (Figure 3D). Interferon-beta therapies (*i.e.,* interferon beta-1a and interferon beta-1b) demonstrated a long-term decline in both cosine similarity and prescription frequency, becoming statistically significant after 2011 [β (95% CI) = –0.019 (–0.030, –0.008), *p*=.002] (Figure 3E). S1P receptor modulators (*i.e.,* fingolimod, siponimod, ozanimod, and ponesimod) showed a non-significant increase in cosine similarity up to 2014 [β (95% CI) = 0.022 (–0.011, 0.055), *p*=.166], followed by a gradual decline [β (95% CI) = –0.026 (–0.043, –0.010), *p*=.005], while prescription frequency rose until around 2015 before decreasing (Figure 3F). Finally, steroids (*e.g.,* cortisone, methylprednisolone, and prednisolone) were frequently used in the management of acute MS relapse and were included as references. Steroids showed a statistically significant decline in cosine similarity prior to 2013 [β (95% CI) = –0.027 (–0.037, –0.018), *p*<.001], followed by a gradual increase [β (95% CI) = 0.029 (0.011, 0.048), *p*=.003], while the prescription frequency consistently increased throughout the follow-up period (Figure 3G).

For completeness, we further examined temporal trends of individual DMT usage among KOMAP-classified MS patients (**Supplementary Figure 1-2** and **Supplementary Table 3**). Among commonly prescribed DMTs, glatiramer acetate and natalizumab exhibited an increasing-to-decreasing pattern, with usage declining after 2011 (*p*<.001 for both). Fingolimod, interferons beta-1a and interferons beta-1b showed no significant trend before 2012 but declined significantly afterward (*p*<.005). Dimethyl fumarate showed a decreasing trend until 2020, while the post-2020 trend was not statistically significant. Ocrelizumab and rituximab displayed increasing trends, with ocrelizumab usage significantly rising after 2018 (*p*=.032), and rituximab significantly increased throughout the follow-up period. In contrast, teriflunomide declined consistently over the follow-up period (*p*=.006). As a reference, methylprednisolone showed a reversal from decreasing to increasing usage after 2014 (*p*=.003). Among less frequently prescribed DMTs (**Supplementary** Figure 3 and **Supplementary Table 3**), cladribine showed an increasing trend until 2019, followed by a non-significant decline. Mitoxantrone consistently decreased in use (*p*=0.013), while alemtuzumab, daclizumab, and peginterferon beta-1a showed no statistically significant trends. Several newer agents (*e.g.,* diroximel fumarate, monomethyl fumarate, ofatumumab, ozanimod, and others) lacked sufficient longitudinal data to evaluate trends.

## Discussion

In this study, we utilized a large, multicenter EHR dataset from two major healthcare systems (UPMC and MGB) spanning both academic and community care settings to evaluate MS diagnostic phenotyping performance using a novel algorithm (*i.e.,* KOMAP) and to examine longitudinal trends in MS therapies through a temporal KG approach. Our findings demonstrate that KOMAP achieved strong performance in identifying MS patients across both cohorts, with higher accuracy observed when using combined codified and NLP-derived feature sets. Notably, our analysis of time-varying relationships between DMTs and MS reveals how clinical practice has shifted over the past two decades in response to emerging therapies and evolving treatment strategies, offering new insight into the dynamics of MS care.

Since the introduction of the first DMT, the management of MS has evolved substantially over the past three decades. Our EHR-based findings from KOMAP-identified MS patients align with these shifting treatment paradigms. Early injectable therapies, such as interferon-beta and glatiramer acetate, were among the first approved DMTs. While moderately effective in reducing relapses, the requirement for frequent injections made long-term adherence challenging.^22,23^ In our study, interferon use remained steady until 2011, then declined significantly. Glatiramer acetate followed a similar trajectory, with a modest increase before 2011 and a subsequent decline, likely due to the emergency of more effective oral therapies.^24^ Oral DMTs, first introduced in the early 2010s, improved convenience and adherence.^25^ Fingolimod and dimethyl fumarate were initially widely adopted, but their use declined over time, possibly due to side effects and the availability of newer options. Teriflunomide also showed a consistent decline, suggesting that many oral agents were eventually supplanted by higher-efficacy treatments. Monoclonal antibodies, such as natalizumab, were initially used for patients with more aggressive disease. Our data showed increased natalizumab use from 2004 to 2011, followed by a decline, consistent with concerns about progressive multifocal leukoencephalopathy (PML).^26^ The most significant shift in MS treatment has been the rise of B-cell depletion therapies, including ocrelizumab, rituximab, and ofatumumab. These therapies have demonstrated strong efficacy in trials and effectiveness in real-world data and are increasingly used as first-line options.^27^ In our analysis, their use began rising around 2014 and accelerated following the approval of ocrelizumab in 2017, reflecting a broader trend toward early, high-efficacy treatment strategies in MS care.

Our study offers several methodological advantages. First, many existing approaches (*e.g.,* calculating feature frequencies across calendar years) fail to capture nuanced relations between DMT prescriptions and disease (*i.e.*, MS). For instance, a high frequency of steroid prescriptions in MS patients does *not* necessarily indicate their primary use for MS treatment, as steroids are also commonly prescribed for various other indications (*e.g.,* allergic reactions, other inflammatory conditions). In contrast, cosine similarity measures the co-occurrence of clinical features, providing a more accurate representation of their relationships.

Second, our method accounts for temporal changes in feature relationships, specifically MS-related DMT prescriptions. The observed prescription trends of MS-related DMTs over time based on the EHR data align with domain knowledge and clinical experience. For instance, the association between natalizumab and MS strengthened after its FDA approval in 2004, peaked several years later, and then gradually declined. Assuming static relationships over time can lead to misleading conclusions, while analyzing the dataset without considering temporal dynamics risks obscuring important trends.

Third, our approach relies solely on summary data (*e.g.,* co-occurrence matrices) to compute cosine similarities and assess relationships trends. This eliminates the need for patient-level data, facilitating cross-institutions collaborations while adhering to privacy regulations. By enabling data sharing, our method expands the scalability of the dataset and enhances the generalizability of findings.

Finally, the novel MS diagnosis algorithm (*i.e.,* KOMAP) enables accurate and efficient identification of MS cases. The AUROC and AUPRC of MS phenotyping exceeded 0.90 in two large, independent healthcare systems. This unsupervised algorithm leverages a publicly available, pre-trained KG of interconnected EHR concepts to efficiently identify informative EHR features, overcoming limitations of expert-selected or less specific feature selection steps. Although the algorithm does not require expert adjudication, the performance metrics reported in this study were validated through manual chart review.

The study also has limitations. First, cosine similarity estimation does not yield a smooth trajectory across calendar years. To address this, we applied curve smoothing to the estimated cosine similarities, which could introduce a discrepancy between the smoothed curve and the constructed confidence intervals. Second, relationships between clinical features may vary not only over time but also with disease progression. For example, the associations between DMT use and MS symptoms may differ depending on whether the patient is in the early or advanced stage of MS. Incorporating disease stages into the analysis (*e.g.,* clustering the EHR data by RRMS and SPMS) could yield more detailed and accurate measurements of feature relationships over time. Third, although the combined dataset includes two large, independent healthcare systems encompassing both academic and community practices, racial and ethnic minorities remain underrepresented. The approach is readily applicable to healthcare systems with more diverse patient populations (*e.g.,* Veterans Affairs Healthcare system) in future studies.

## Conclusion

In summary, our study demonstrates the effectiveness of advanced phenotyping algorithm KOMAP for accurately and efficiently identifying MS patients from EHR data across two large healthcare systems and highlights the value of the temporal knowledge graph approach in capturing real-world treatment dynamics of a chronic neurological condition.

## Data Availability

Code for analysis and figures is available at <https://github.com/xialab2016/MSphenotypingandDMTtrend.git>. We will publicly disseminate anonymous summary-level registry data and EHR data. The rationale for not sharing patient-level data is that patient-level clinical data (either de-identified information or limited protected health information containing dates of clinical events or even if anonymous due to concern for re-identification) are universally subject to the rules and regulation of each healthcare system, which may only be affiliated with but are not the same as the primary academic institutions of the study investigators. Sharing of de-identified EHR data with qualified external researchers by each of the study performance site may be permissible only after the approval of the respective Institutional Review Boards (IRBs), regulatory oversight agents of the healthcare systems (that own the clinical data) as well as the appropriate Data Usage Agreements (DUA) between institutions.

## Acknowledgment

The authors thank all research participants and the clinicians from UPMC and MGB.

## Funding Source

This study was funded by the National Institute of Neurological Disorders and Stroke of the National Institutes of Health under award numbers R01 NS098023 (Z. Xia).

## Conflict of Interest

Not applicable.

## Supplementary Tables and Figures

**S-Figure 1.**
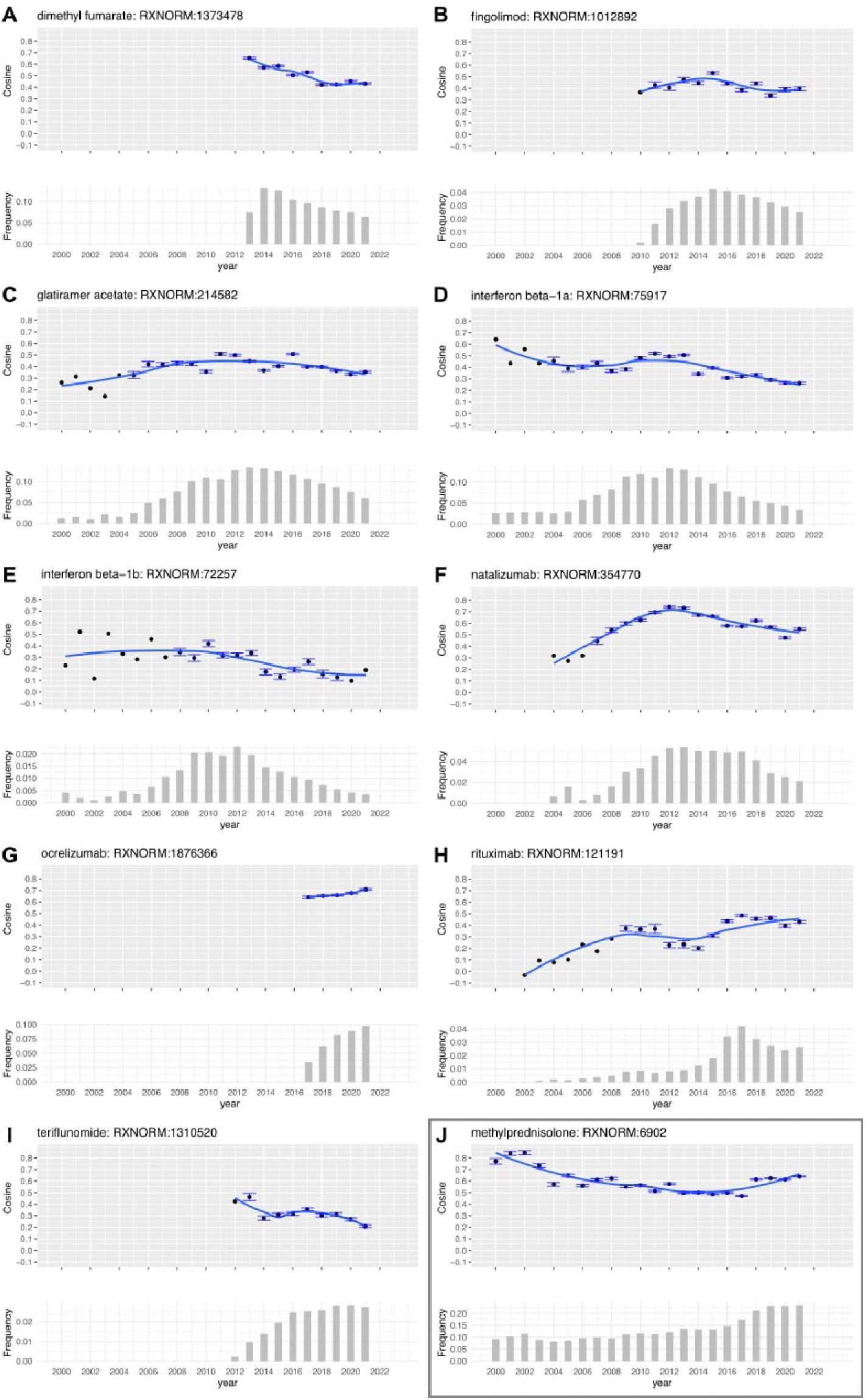
Trends of commonly prescribed MS-related DMTs (2004-2022) **A.** dimethyl fumarate, **B.** fingolimod, **C.** glatiramer acetate, **D.** interferon beta-1a, **E.** interferon beta-1b, **F.** natalizumab, **G.** ocrelizumab, **H.** rituximab, **I.** terifluonomide, and **J.** methylprednisolone (included as a reference). For each DMT, the top panel displays the dynamic cosine similarity with 95% confidence intervals. The bottom panel shows the DMT frequency, defined as the proportion of MS patients prescribed the target drug in each calendar year.

**S-Figure 2.**
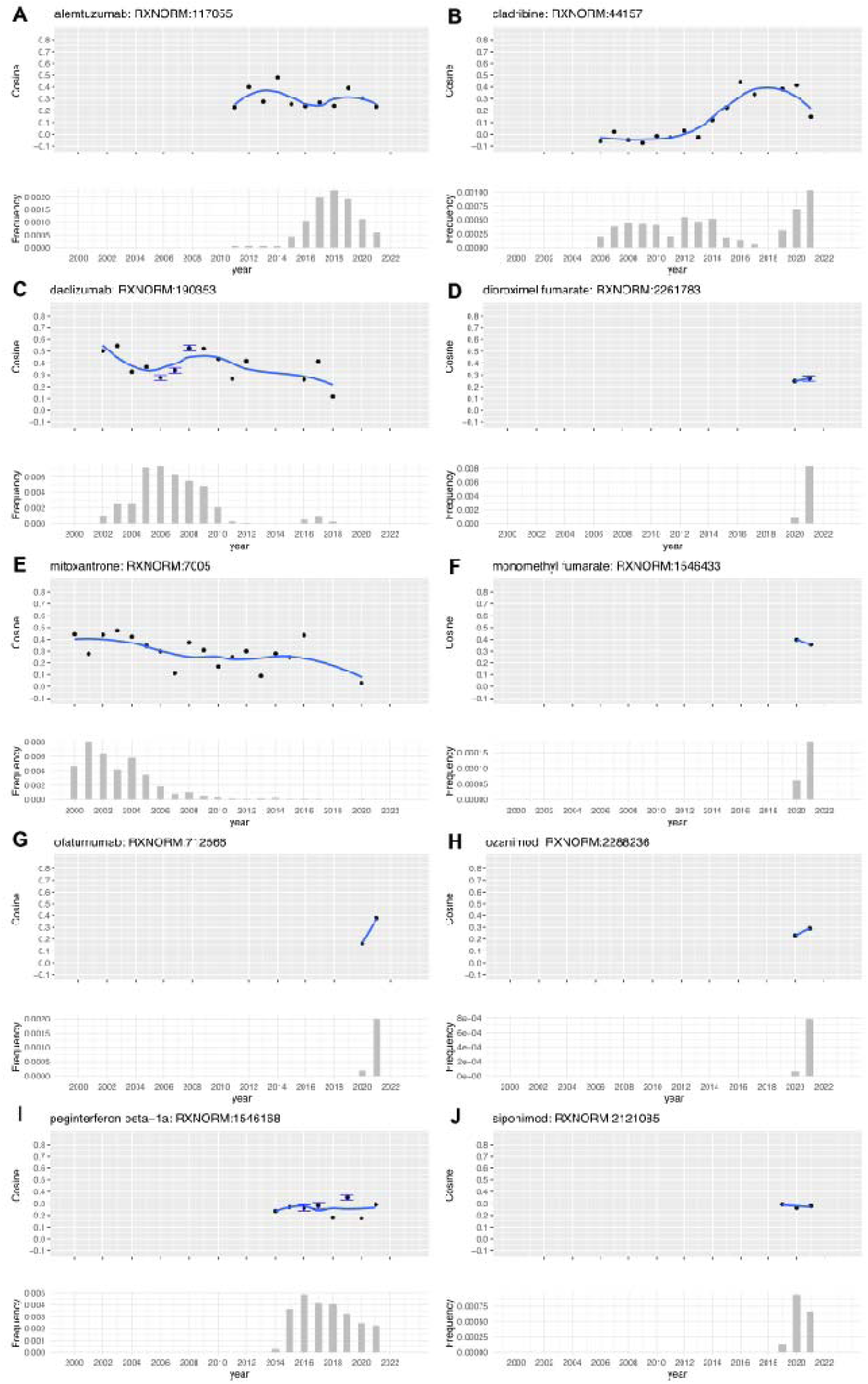
Trends of less commonly prescribed MS-related DMTs (2004 and 2022). **A.** alemtuzumab, **B.** cladribine, (**C.** daclizumab (withdrawn), **D.** dioroximel fumarate, **E.** mitoxantrone, **F.** monomethyl fumarate, **G.** ofatumumab, **H.** ozanimod, **I.** peginterferon beta-1a and **J.** siponimod. For each DMT, the top panel displays the dynamic cosine similarity with 95% confidence intervals. The bottom panel shows the DMT frequency, defined as the proportion of MS patients prescribed the target drug in each calendar year.

---

**S-Table 1.** MS phenotyping model (KOMAP) features and coefficients.

| Feature<sup>1</sup> | Description | UPMC | MGB |
| --- | --- | --- | --- |
|  |  | Coefficient | Coefficient |
| C0004093 | Asthenia | 0.006 | 0.000 |
| C0004268 | Attention | 0.000 | -0.013 |
| C0010957 | Tissue damage | 0.000 | 0.000 |
| C0011303 | Demyelinating Diseases | 0.003 | 0.020 |
| C0011304 | Demyelination | 0.005 | 0.019 |
| C0011570 | Mental Depression | 0.000 | 0.000 |
| C0011581 | Depressive disorder | -0.002 | 0.000 |
| C0013415 | Dysthymic Disorder | 0.000 | 0.000 |
| C0026769 | Multiple Sclerosis | 0.573 | 0.545 |
| C0026838 | Muscle Spasticity | 0.023 | 0.038 |
| C0029134 | Optic Neuritis | 0.009 | 0.023 |
| C0035020 | Relapse | 0.031 | 0.054 |
| C0037179 | Unmarried person | -0.001 | -0.009 |
| C0041696 | Unipolar Depression | 0.000 | 0.000 |
| C0066677 | modafinil | 0.000 | 0.000 |
| C0069426 | Oligoclonal Bands (protein) | 0.000 | 0.000 |
| C0087136 | Unmarried | -0.001 | -0.009 |
| C0159028 | General symptom | 0.000 | 0.000 |
| C0163712 | Relate - vinyl resin | -0.002 | -0.009 |
| C0202205 | Oligoclonal protein measurement | 0.000 | 0.000 |
| C0231170 | Disability | 0.001 | 0.000 |
| C0289884 | glatiramer acetate | 0.000 | 0.000 |
| C0344315 | Depressed mood | 0.000 | 0.000 |
| C0349217 | Depressive episode, unspecified | 0.000 | 0.000 |
| C0423551 | Sensory symptoms | 0.000 | 0.000 |
| C0528175 | Copaxone | 0.040 | 0.065 |
| C0682148 | Disability status | 0.000 | 0.000 |
| C0722882 | Provigil | 0.009 | 0.016 |
| C0751967 | Multiple Sclerosis, Relapsing-Remitting | 0.018 | 0.035 |
| C0878575 | Peripheral demyelination | 0.005 | 0.020 |
| C1172734 | natalizumab | 0.000 | 0.000 |
| C1269683 | Major Depressive Disorder | 0.000 | 0.000 |
| C1457887 | Symptoms | 0.000 | 0.000 |
| C1529600 | Tysabri | 0.035 | 0.014 |
| C1579931 | Depressed - symptom | 0.000 | 0.000 |
| C1857639 | Dysmyelination | 0.000 | 0.000 |
| C2362914 | Clinical depression | 0.000 | 0.000 |
| C2752009 | White matter lesion | 0.004 | 0.035 |
| C3540014 | Central Nervous System Diagnostic Radiopharmaceuticals | 0.000 | 0.000 |
| C3714552 | Weakness | 0.004 | 0.000 |
| C5202921 | RECIL PD | 0.000 | 0.000 |
| C5203119 | Intensity and Distress 5 | -0.014 | -0.008 |
| PheCode:296 | mood disorders | 0.000 | 0.000 |
| PheCode:296.2 | depression | 0.000 | 0.000 |
| PheCode:296.22 | major depressive disorder | 0.000 | 0.000 |
| PheCode:300 | anxiety disorders | -0.004 | 0.000 |
| PheCode:334 | degenerative disease of the spinal cord | 0.000 | 0.000 |
| PheCode:335 | multiple sclerosis | 0.584 | 0.648 |
| PheCode:341 | other demyelinating diseases of central nervous system | 0.000 | 0.000 |
| PheCode:350.1 | abnormal involuntary movements | 0.000 | 0.000 |
| PheCode:377.3 | optic neuritis/neuropathy | 0.000 | 0.000 |
| PheCode:596.5 | functional disorders of bladder | 0.023 | 0.016 |
| RXNORM:1012892 | fingolimod | 0.003 | 0.000 |
| RXNORM:1292 | baclofen | 0.019 | 0.014 |
| RXNORM:1310520 | teriflunomide | 0.000 | 0.000 |
| RXNORM:1373478 | dimethyl fumarate | 0.027 | 0.012 |
| RXNORM:214582 | glatiramer | 0.011 | 0.002 |
| RXNORM:30125 | modafinil | 0.010 | 0.004 |
| RXNORM:354770 | natalizumab | 0.024 | 0.000 |
| RXNORM:72257 | interferon beta-1b | 0.000 | 0.000 |
| RXNORM:75917 | interferon beta-1a | 0.035 | 0.009 |
| RXNORM:897018 | dalfampridine | 0.011 | 0.000 |
| utls | Healthcare Utility | -0.542 | -0.200 |

<sup>1</sup> Feature: We used the Online Narrative and Codified feature Search engine (*i.e.*, ONCE) to select 20 codified and 47 NLP-derived features relevant for MS.

**S-Table 2.** MS phenotyping model (KOMAP) performance with different feature sets.

|  |  | UPMC |  |  |  | MGB |  |  |  |
| --- | --- | --- | --- | --- | --- | --- | --- | --- | --- |
|  |  | Main PheCode<sup>1</sup> | Main CUI<sup>2</sup> | Codified<sup>3</sup> | Both<sup>4</sup> | Main PheCode<sup>1</sup> | Main CUI<sup>2</sup> | Codified<sup>3</sup> | Both<sup>4</sup> |
| **AUROC** |  | 0.854 | 0.798 | 0.912 | 0.922 | 0.942 | 0.983 | 0.974 | 0.994 |
| **AUPROC** |  | 0.941 | 0.898 | 0.963 | 0.966 | 0.831 | 0.839 | 0.879 | 0.94 |
| **0.970 Specificity** | **Sensitivity** | 0.502 | 0.209 | 0.631 | 0.674 | 0.605 | 0.747 | 0.609 | 0.957 |
|  | **PPV** | 0.976 | 0.944 | 0.981 | 0.982 | 0.876 | 0.897 | 0.868 | 0.912 |
|  | **NPV** | 0.436 | 0.327 | 0.510 | 0.541 | 0.881 | 0.921 | 0.884 | 0.986 |
| **0.950 Specificity** | **Sensitivity** | 0.545 | 0.268 | 0.711 | 0.698 | 0.637 | 0.82 | 0.645 | 0.957 |
|  | **PPV** | 0.967 | 0.935 | 0.975 | 0.974 | 0.876 | 0.847 | 0.808 | 0.861 |
|  | **NPV** | 0.454 | 0.341 | 0.567 | 0.557 | 0.888 | 0.941 | 0.892 | 0.985 |
| **0.900 Specificity** | **Sensitivity** | 0.662 | 0.425 | 0.791 | 0.794 | 0.716 | 0.921 | 1.000 | 1.000 |
|  | **PPV** | 0.951 | 0.92 | 0.952 | 0.953 | 0.732 | 0.761 | 0.764 | 0.764 |
|  | **NPV** | 0.518 | 0.385 | 0.630 | 0.634 | 0.907 | 0.971 | 1.000 | 1.000 |

<sup>1</sup> Main MS PheCode: KOMAP with log(1+ aggregated MS PheCode counts).
<sup>2</sup> Main MS CUI: KOMAP with log(1+ aggregated MS CUI counts).
<sup>3</sup> Codified: KOMAP with log(ICD+1) as surrogate plus 20 codified features selected by ONCE.
<sup>4</sup> Both: KOMAP with log(ICD+1) and log(NLP+1) as surrogate plus 20 ONCE-selected codified features plus 47 ONCE-selected NLP-derived narrative features.
*ONCE*, Online Narrative and Codified feature Search engine powered by the Online Narrative and Codified feature Search engine

**S-Table 3.** Trends of individual MS-related DMT between 2004 and 2022.

| Common Prescribed DMTs |  |  |  |  |  |  |  |
| --- | --- | --- | --- | --- | --- | --- | --- |
| DMT Class | Follow-up Period | Trend<sup>1</sup> | Change Point | Before Change Point |  | After Change Point |  |
| | | | | $\beta$ (95% CI) | p | $\beta$ (95% CI) | p |
| dimethyl fumarate | 2013-2022 | D-NA | 2020 | -0.027 (-0.042, -0.012) | 0.004 | -0.091 (-0.229, 0.046) | 0.155 |
| fingolimod | 2010-2022 | NA-D | 2012 | 0.022 (-0.010, 0.053) | 0.156 | -0.026 (-0.042, -0.011) | 0.004 |
| glatiramer acetate | 2000-2022 | I-D | 2011 | 0.22 (0.012, 0.032) | <0.001 | -0.013 (-0.025, -0.002) | 0.026 |
| interferon beta-1a | 2000-2022 | NA-D | 2012 | -0.006 (-0.016, 0.005) | 0.262 | -0.022 (-0.037, -0.007) | 0.007 |
| interferon beta-1b | 2000-2022 | NA-D | 2002 | 0.036 (-0.117, 0.190) | 0.625 | -0.013 (-0.021, -0.004) | 0.005 |
| natalizumab | 2004-2022 | I-D | 2011 | 0.063 (0.050, 0.077) | <0.001 | -0.027 (-0.035, -0.019) | <0.001 |
| ocrelizumab | 2017-2022 | NA-I | 2018 | 0.010 (-0.039, 0.059) | 0.465 | 0.020 (0.004, 0.035) | 0.032 |
| rituximab | 2022-2022 | I-I | 2008 | 0.045 (0.016, 0.074) | 0.004 | 0.012 (0.002, 0.023) | 0.020 |
| teriflunomide | 2012-2022 | D | NA | -0.017 (-0.027, -0.006) | 0.006 | -0.017 (-0.027, -0.006) | 0.006 |
| methylprednisolone | 2000-2022 | D-I | 2014 | -0.023 (-0.029, -0.016) | <0.001 | 0.029 (0.011, 0.046) | 0.003 |
| Less Prescribed DMTs |  |  |  |  |  |  |  |
| alemtuzumab | 2011-2022 | NA | NA | -0.011 (-0.028, 0.007) | 0.207 | -0.011 (-0.028, 0.007) | 0.207 |
| cladribine | 2006-2022 | I-NA | 2019 | 0.039 (0.024, 0.055) | <0.001 | -0.106 (-0.261, 0.048) | 0.159 |
| daclizumab | 2002-2018 | NA | NA | -0.012 (-0.025, 0.001) | 0.064 | -0.012 (-0.025, 0.001) | 0.064 |
| dioroximel fumarate | 2020-2022 | NA | NA | NA | NA | NA | NA |
| mitoxantrone | 2000-2020 | D | NA | -0.013 (-0.023, -0.003) | 0.013 | -0.013 (-0.023, -0.003) | 0.013 |
| monomethyl fumarate | 2020-2022 | NA | NA | NA | NA | NA | NA |
| ofatumumab | 2020-2022 | NA | NA | NA | NA | NA | NA |
| ozanimod | 2020-2022 | NA | NA | NA | NA | NA | NA |
| peginterferon beta-1a | 2014-2022 | NA | NA | 0.008 (-0.012, 0.028) | 0.383 | 0.008 (-0.012, 0.028) | 0.383 |

<sup>1</sup> **Trend:** the pattern of MS-related DMT prescription over time assessed using cosine similarity.
I, increasing throughout;
I-D, increasing trend before the change point, followed by a decreasing trend;
I-I, increase throughout, at different rate before and after the change point;
I-NA, increasing trend with no significant trend after the change point;
D, decreasing throughout;
D-I, decreasing trend, followed by an increasing trend;
D-NA, decreasing trend with no significant trend after the change point;
NA-D, no significant trend before the change point, followed by a decreasing trend;
NA-I, no significant trend before the change point, followed by an increasing trend;

## Notes

### Competing Interest Statement

The authors have declared no competing interest.

